# Figure format and the accuracy of AI-assisted reconstruction of Kaplan–Meier survival data

**DOI:** 10.64898/2026.11.03.26367147

**Authors:** Shiyin Li, Xing Xing, Bulun Te, Han Han, Haoyu Ji, Yulin Yu, Lianne Siegel, Lifeng Lin

## Abstract

Reconstructing participant-level data from published Kaplan–Meier curves supports meta-analysis and other secondary survival analyses when original trial records are unavailable. Researchers may use original PDF figures or screenshots, which can look alike but retain different information. We assessed how converting vector figures to images affects reconstruction by a language model that can inspect files and execute code.

We generated 64 independent two-arm datasets across four scenarios combining 100 or 1,000 participants per arm with proportional or crossing hazards. Each figure was supplied as a native vector PDF and a matched 300-dpi PNG to Opus 5.5 through Claude Code under the same instructions. Outcomes compared reconstructed records with the original observed records: time-averaged absolute curve error through 24 months, error in the between-group restricted mean survival time difference, and error in the Cox log-hazard ratio.

Native PDFs had lower curve error in all 64 completed pairs, including replacements for incomplete runs. Mean errors were 0.000029 and 0.1047 percentage points for PDF and PNG, respectively; the paired difference was −0.1047 (95% confidence interval, −0.1194 to −0.0909).

PNG errors in the restricted mean survival time difference and log-hazard ratio were small: mean absolute errors were 0.0084 months and 0.0027. Lower resolution and compression increased curve errors; embedding unchanged pixels in a PDF did not restore vector accuracy. Some PNG runs exceeded the instructed revision limit, so results describe the implemented workflow rather than performance under a strictly enforced limit.

Preserving vector information improved reconstruction fidelity in this workflow, but the observed differences do not establish clinically consequential changes in treatment estimates. Validation against reported survival summaries and event counts remains necessary. Generalization to published clinical figures and other model workflows requires further evaluation.

**Author summary:** Clinical studies often publish survival graphs without sharing the underlying patient records. Researchers can estimate event and follow-up times from these graphs and use them to combine evidence across studies. Artificial intelligence may reduce the work involved, but its accuracy can depend on how a figure is supplied.

We studied whether an original vector PDF and a matching image led the same artificial intelligence system to reconstruct different data. Using simulated studies allowed us to compare every reconstruction with known records. The system could write and run computer code. It recovered curves more accurately from vector PDFs, which preserved readable curve coordinates, than from images containing only pixels. However, average differences in treatment estimates were small. Lower-resolution and compressed images produced larger errors, and putting an image inside a PDF did not recover the lost information.

Our findings support preserving original vector figures when possible. Even when reconstructed KM curves closely matched the originals, the reconstructed data sometimes contained incorrect event counts. The results apply to one computational workflow using clean simulated figures; performance with published clinical figures, other systems, and consistently enforced revision limits still needs evaluation.

**Data availability statement:** All data used in this study were simulated. The project repository is https://github.com/ FionaShiyinLi/km-curve-reconstruction-study. The reproducibility materials comprise reference records, input figures, reconstructed records, dataset-level results, reconstruction instructions, random seeds, and generation, reconstruction, and analysis code.

## 1 Introduction

Independent evaluation and reuse of clinical trial findings can be difficult when original individual participant data (IPD) are unavailable, partly because of privacy concerns and data-sharing restrictions [1]. Generative AI may help researchers extract and reconstruct quantitative information from published reports for secondary analyses and evidence synthesis [2, 3]. However, the reliability of AI-assisted reconstruction may depend on how information is preserved in the source figures.

For survival outcomes, published Kaplan–Meier (KM) curves are often an important source of information when original IPD are unavailable [4]. In a systematic review of randomized cancer trials, 86 of 98 reports presented an overall-survival KM curve in the main article [5]. Reconstructing approximate event and censoring records from these curves can support meta-analysis and secondary analyses of survival outcomes [6, 7]. These records can reproduce marginal survival summaries and treatment comparisons, but do not recover unreported participant covariates or provide all the advantages of original IPD.

Conventional reconstruction usually separates curve digitization from statistical estimation of event and censoring times [8]. The Guyot algorithm reconstructs participant-level event and censoring times from digitized KM curve coordinates and reported numbers at risk, using total event counts when available [7]. Rogula et al. incorporated marked censoring times, and Titman formulated reconstruction as a constrained optimization problem [9, 10]. Preparing inputs for these methods can require substantial manual work, including curve digitization and transcription of numbers at risk. Automated tools such as SurvdigitizeR can reduce the effort needed to extract curve coordinates from images [11].

Recent systems have begun using multimodal large language models (LLMs) for KM reconstruction. KM-GPT combines image processing, multimodal interpretation, and iterative reconstruction. In simulations, it successfully processed 538 of 540 KM plots and achieved a median integrated absolute curve error of 0.018 among successful cases [2]. KMGen uses an LLM-guided code-writing agent for curve extraction and reported a mean integrated absolute error of 0.0151 across 32 simulated plots, including challenging layouts [3]. General-purpose models can also inspect figures and write executable reconstruction code when equipped with appropriate tools [12, 13]. However, these studies do not establish how figure format affects reconstruction accuracy when the underlying KM figure is unchanged.

A native vector PDF may preserve curve paths, text, and censoring marks as geometric objects, whereas a screenshot or PNG contains pixels [14, 15]. Researchers nevertheless commonly use images extracted from articles: Kim et al. reconstructed data from screenshots of 46 KM figures across 14 oncology articles [16]. The distinction is therefore relevant to evidence synthesis. RESOLVE-IPD already exploits vector coordinates and censoring marks in a specialized reconstruction workflow [17]. However, this does not quantify what happens when the same figure is converted to pixels before processing by a general-purpose model.

Everest et al. digitized published KM curves from 39 cancer trials using DigitizeIt and reconstructed individual participant data using the Guyot algorithm. They compared RMST estimates with those from the original trial data and found that reconstruction accuracy was generally not associated with whether the published figures were vector-traced or bitmapped [18]. A paired experiment on different representations of the same figure addresses a complementary question by holding the underlying data and layout fixed. We used a general-purpose, tool-enabled LLM (Opus 5.5 via Claude Code) to reconstruct individual participant data from KM figures. The model could inspect figures and write and execute reconstruction code, without access to the original participant records or dedicated KM reconstruction software. For each of 64 simulated datasets, we supplied the same KM figure as a native vector PDF and a matched 300-dpi PNG under identical instructions. We compared the reconstructed KM curves and treatment-effect estimates with those from the original records. Exploratory analyses examined censoring marks, image quality, plotting style, and a separate automated digitization–Guyot workflow.

## 2 Methods

### 2.1 Study design and simulated survival data

We conducted a paired simulation study of 64 independent two-arm datasets [19]. Each dataset was presented to the model as a native vector PDF and as a matched PNG rendered at 300 dpi (PNG300; 1,800 × 1,350 pixels), giving 128 planned primary reconstructions. We used a balanced 2 × 2 design, with 16 datasets in each combination of sample size (100 or 1,000 participants per arm) and hazard pattern (proportional or crossing hazards; Fig 1 and Table 1).

**Fig 1.**
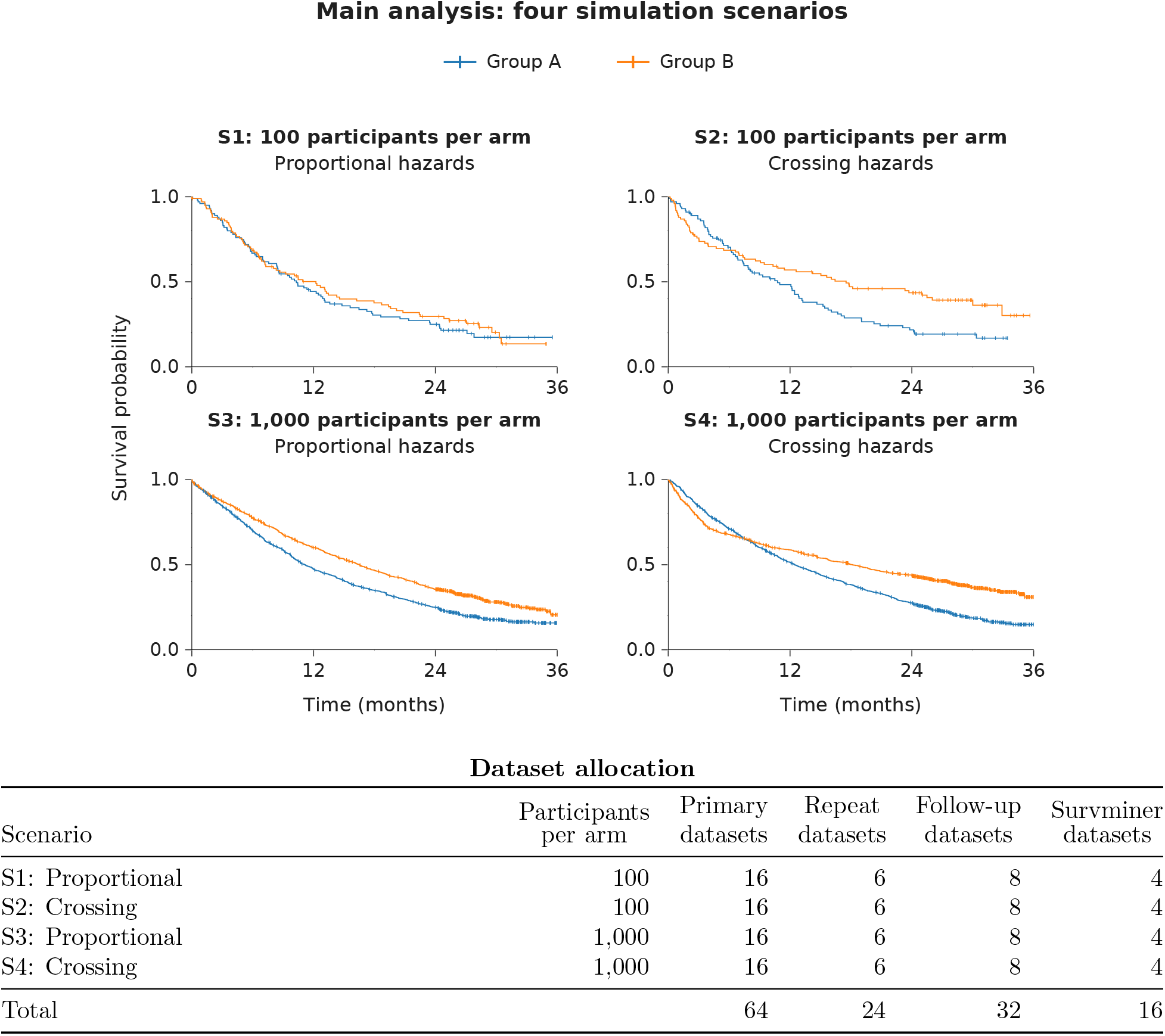
Simulation scenarios, example input curves, and dataset allocation. The four panels show example KM curves for the four simulation scenarios. Counts below refer to datasets. Each primary dataset supplied one native PDF and one PNG300; each repeat dataset supplied one additional run per format.

**Table 1.** Overview of the main format comparison and exploratory analyses. Counts refer to independent datasets

| Analysis | Comparison | Datasets |
|---|---|---|
| **Main format comparison** |  |  |
| File format | Native vector PDF vs PNG300 | 64 |
| **Exploratory analyses** |  |  |
| Censoring marks | PDF without marks vs PDF; PNG300 without marks vs PNG300 | 32 |
| Image resolution | PNG150 and PNG100, each vs PNG300 | 32 |
| JPEG compression | JPEG100 (quality 75) vs PNG100 | 32 |
| Vector content | Image-only PDF vs PNG300 | 32 |
| Plotting style | survminer-generated plots: native PDF vs PNG300 | 16 |
| Automated Guyot reconstruction workflow | Automated digitization plus Guyot vs LLM, using the same PNG300 figures | 64 |

The datasets were generated in two batches of 32, with eight datasets per scenario in each batch. All 32-dataset exploratory analyses used the first batch. The plotting-style comparison used 16 datasets randomly selected from this batch (four per scenario). The automated digitization–Guyot comparison used all 64 datasets.

Sample size was varied because larger groups produce more numerous, smaller KM steps at a fixed plotting resolution. Proportional and crossing-hazards scenarios represented distinct patterns encountered in survival analyses [20, 21]. For group A, event times followed an exponential distribution with monthly hazard *λ_A_* = log(2)*/*12, giving a median of 12 months. Group B had hazards

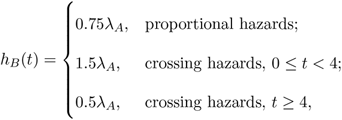

where *t* is time since enrollment in months. The crossing-hazards specification produces theoretical survival curves that cross at 8 months. The generating distributions were exponential or piecewise exponential; no other event-time distributions were evaluated.

Enrollment time *E_i_* was uniform on 0–12 months, dropout time *D_i_* was exponential with monthly rate 0.005, and administrative censoring occurred at *C_i_* = 36 − *E_i_*. Event, enrollment, and dropout times were generated independently. For event time *T_i_*, the observed record was

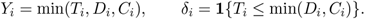

Thus, administrative censoring occurred between 24 and 36 months, while random dropout could occur earlier. Reference estimates were calculated from these observed records, rather than from the generating survival distributions. The datasets were generated in two independent batches of 32 with dataset-specific random-number streams; seeds and generation details are given in S1 Text [19, 22].

For each simulated dataset, we generated a KM figure showing both survival curves, censoring marks, and numbers at risk at 6-month intervals through month 36. We used ReportLab to create a 6-by-4.5-inch vector PDF and then converted it to a 1,800-by-1,350-pixel PNG (300 dpi) using Poppler. Both formats displayed the same information, but only the native PDF preserved the original vector coordinates and text.

### 2.2 Reconstruction procedure

All LLM reconstructions used Opus 5.5 (claude-opus-5-5; Anthropic) through Claude Code 2.1.280, with medium reasoning effort and identical instructions. In a preliminary test of eight figure inputs from four datasets, medium effort successfully generated complete participant-level datasets for all eight inputs, compared with seven using high effort.

Each reconstruction began in a fresh session with one figure, group sizes, time units, and a seed for any stochastic code. The model could inspect files and write and execute code using general-purpose tools, but could not use dedicated KM-to-IPD software, simulated participant-level data, or pre-extracted coordinates. It returned group, observed follow-up time, and event status for each reconstructed participant and checked these against the figure and risk table. Run order was randomized across scenarios and formats.

Sessions had a 60-minute time limit and the same nominal resource settings. Instructions allowed two revisions after the first complete reconstruction, but this limit was not automatically enforced. The final analysis retained complete outputs from sessions exceeding the limit and included replacement runs for incomplete outputs; failures and departures from the instructed procedure are reported below and in S1 Text. Accordingly, the comparison evaluates the implemented workflow, including its retry policy, rather than performance under a strictly enforced revision limit.

Repeatability was assessed in 24 prespecified datasets by one additional reconstruction per format. Differences between first and repeated curves were summarized separately from errors relative to the reference records; repeats did not increase the number of independent datasets.

### 2.3 Accuracy outcomes

The reference for each outcome was the estimate from the original observed records. The primary outcome was time-averaged absolute KM error (TAAE) through *τ* = 24 months, averaged across groups:

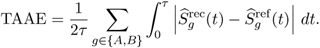

The integral was calculated exactly between right-continuous steps. TAAE is on the survival-probability scale in this expression; we multiply it by 100 when reporting percentage points (pp). We evaluated KM curve error through month 24, before administrative censoring began, to compare reconstructions over a common period with relatively well-supported risk sets. We also evaluated KM curve accuracy beyond month 24 (S12 Table in S1 Text).

Secondary outcomes measured absolute errors in the between-group RMST difference through 24 months and the Cox log-hazard ratio [23, 24]. Let *k* ∈ {rec, ref} denote the reconstructed and reference datasets, respectively. The RMST contrast was

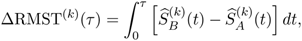

and its error was |ΔRMST^(rec)^ − ΔRMST^(ref)^|. Cox models used full observed follow-up and Efron’s tie approximation; error was |*β*^rec^ − *β*^ref^ |, with *β* denoting the group B versus A log-hazard ratio [25–27]. Under crossing hazards, this evaluates reproduction of the empirical Cox estimate, not estimation of a constant underlying hazard ratio.

Exploratory outcomes included maximum absolute pointwise curve error, total event counts, and estimated standard errors. Full-follow-up event counts and Cox estimates use a longer window than TAAE and RMST; agreement across these outcomes therefore requires cautious interpretation. Scoring conventions and additional outcomes are specified in S1 Text.

We conducted a post hoc analysis of event counts and KM curve accuracy in the censoring-mark experiment. We compared reconstructed and reference event counts separately through month 24 and after month 24. We also calculated KM curve error from month 24 to the earliest last-observed time across both reference arms and all four reconstruction conditions, capped at month 36 (S12 Table in S1 Text).

### 2.4 Statistical analysis

We calculated the mean paired difference in reconstruction error between native PDF and PNG300, with equal weighting across the four scenarios. Negative differences favor PDF. To obtain 95% confidence intervals (CIs), we used 10,000 stratified paired bootstrap samples of the existing reconstruction results. In each bootstrap sample, we resampled 16 datasets with replacement within each scenario, keeping the PDF and PNG300 results for each dataset paired. We then recalculated the mean difference. These CIs quantify uncertainty in the average PDF–PNG300 difference across datasets generated under the four simulation scenarios [28]. The same procedure was applied to secondary outcomes.

### 2.5 Exploratory analyses

We examined how figure characteristics affected reconstruction accuracy using the first eight of the 16 datasets generated in each of the four simulation scenarios (32 datasets in total), referred to as the *32-dataset exploratory subset*. We examined the effects of censoring marks, image resolution, and compression by creating five modified versions of each KM figure: native PDF and PNG300 without censoring marks, PNGs at 150 and 100 dpi, and a quality-75 JPEG derived from the 100-dpi PNG, yielding 160 additional reconstructions. Removing censoring marks preserved the curves and numbers at risk. Model settings, instructions, and computational limits were held constant, and run order was randomized.

Each modified input was compared with its corresponding reference: the original PDF for the no-mark PDF, the original PNG300 for the no-mark PNG300 and lower-resolution PNGs, and the 100-dpi PNG for JPEG compression. Original PDF and PNG300 results were reused from the primary experiment, whereas modified inputs were processed in later sessions. These exploratory contrasts may therefore also reflect differences between execution periods. For each comparison, we calculated the mean paired TAAE difference (modified minus reference) and its 95% confidence interval using the scenario-stratified bootstrap procedure described above.

In a comparison specified after the LLM results were known, we evaluated an automated digitization–Guyot workflow on the same 64 PNG300 figures. For each figure, a fixed image-processing pipeline traced the KM curves using predefined color thresholds and used optical character recognition (OCR) to read numeric axis labels, group labels, and numbers at risk. The pipeline did not use an LLM, manual point selection, or figure-specific correction.

We then applied our implementation of the Guyot algorithm to the extracted coordinates and risk-table information to estimate event counts and distribute censoring times between reported risk-table time points [7]. The workflow had no access to the original participant-level records or native PDF coordinates, and the Guyot reconstruction did not use total event counts or visible censoring marks.

In a post hoc validation, our numerical implementation matched the published Guyot R code for all 256 arm-level inputs when both received identical prepared coordinates, risk-table interval indices, and numbers at risk (S1 Text). We compared this automated digitization–Guyot workflow with the LLM workflow using the same accuracy outcomes and reference data.

To assess whether the PDF–PNG300 accuracy difference persisted with a different plotting style, we redrew the KM figures for 16 datasets (four per scenario) using survminer, an R package for creating publication-ready survival plots with risk tables [29]. These datasets were randomly selected from the 32-dataset exploratory subset without considering reconstruction accuracy. These 16 datasets provided 32 new reconstructions, one per format. We kept the underlying KM data, risk tables, censoring marks, figure dimensions, and axis ranges unchanged, along with the reconstruction settings.

A further experiment placed the unchanged PNG300 image inside a PDF for all 32 follow-up datasets. These image-only PDFs retained the pixels but had no separately readable curve coordinates or text, allowing us to examine the role of vector content beyond the PDF file type (S1 Text).

## 3 Results

### 3.1 Completion and primary reconstruction accuracy

Two primary PNG300 sessions did not produce participant records and were later replaced. One native-PDF assignment failed before any model request and was also replaced. Before the two PNG replacements, 62 pairs were complete; the final analysis included 64 pairs. Among the initial 62 completed pairs, 12 PNG sessions exceeded the instructed two-revision limit. In a post hoc analysis excluding these pairs, the mean TAAE difference in the remaining 50 pairs was −0.0943 pp (95% CI, −0.1068 to −0.0833). This supports the direction of the format difference but does not recreate a comparison under an enforced revision limit (S6 Table in S1 Text).

Native PDFs reconstructed the reference KM curves nearly exactly, whereas PNG300 images introduced small but consistently larger errors. Mean TAAE was 2.91 × 10^−5^ pp for native PDF and 0.1047 pp for PNG300; the PNG300 median was 0.093 pp (interquartile range, 0.064–0.118). Native PDFs had lower TAAE than PNG300 images in all 64 paired datasets (mean paired difference, −0.1047; 95% CI, −0.1194 to −0.0909; Table 2, Fig 2). An example illustrates the small visible differences despite measurable reconstruction errors (Fig 3). The PDF advantage was consistent across all four simulation scenarios. Scenario-specific mean paired differences ranged from −0.1094 to −0.0988 pp, with all four 95% confidence intervals below zero (S1 Table in S1 Text).

**Fig 2.**
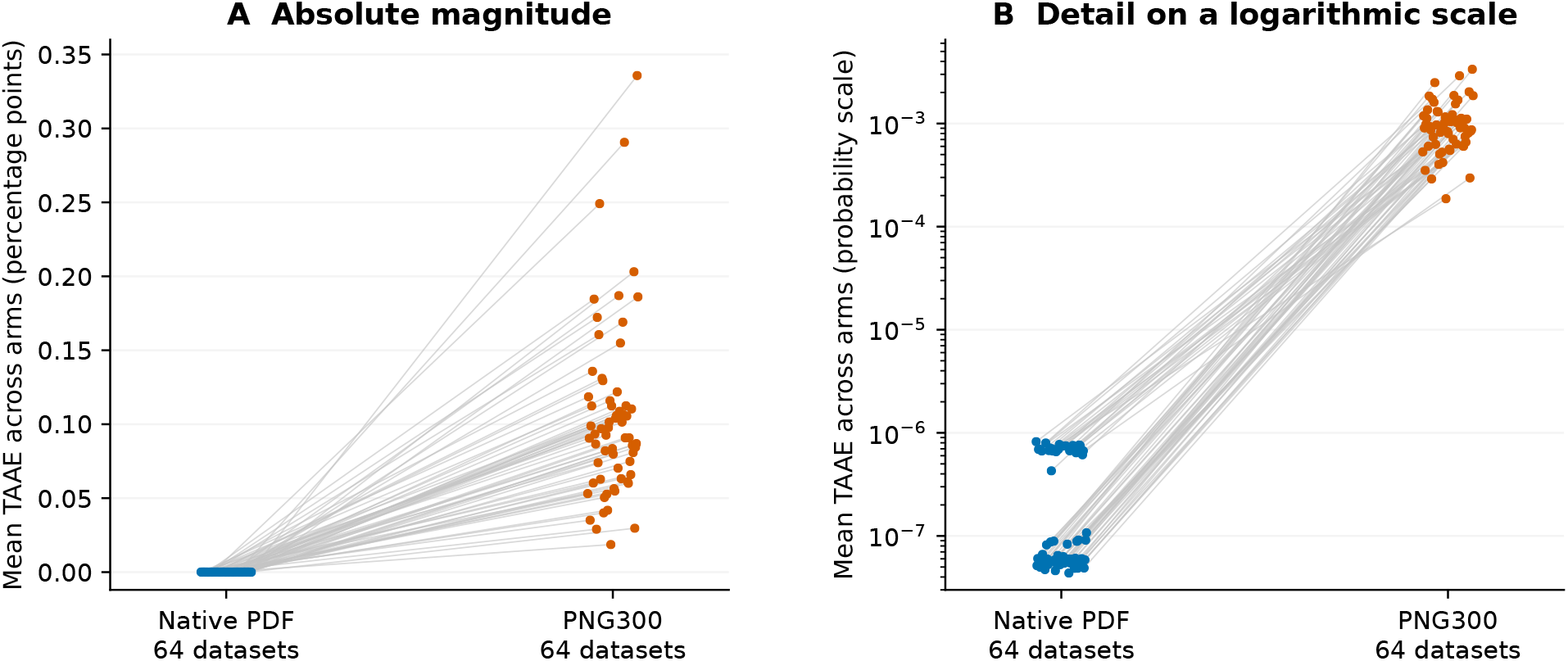
Primary reconstruction error in 64 independent datasets. Native PDF and PNG300 results are paired within each dataset. TAAE averages absolute curve differences through 24 months across both groups. The panel labels specify the scale; probability-scale values are multiplied by 100 to obtain percentage points. Negative PDF-minus-PNG300 differences favor native PDF.

**Fig 3.**
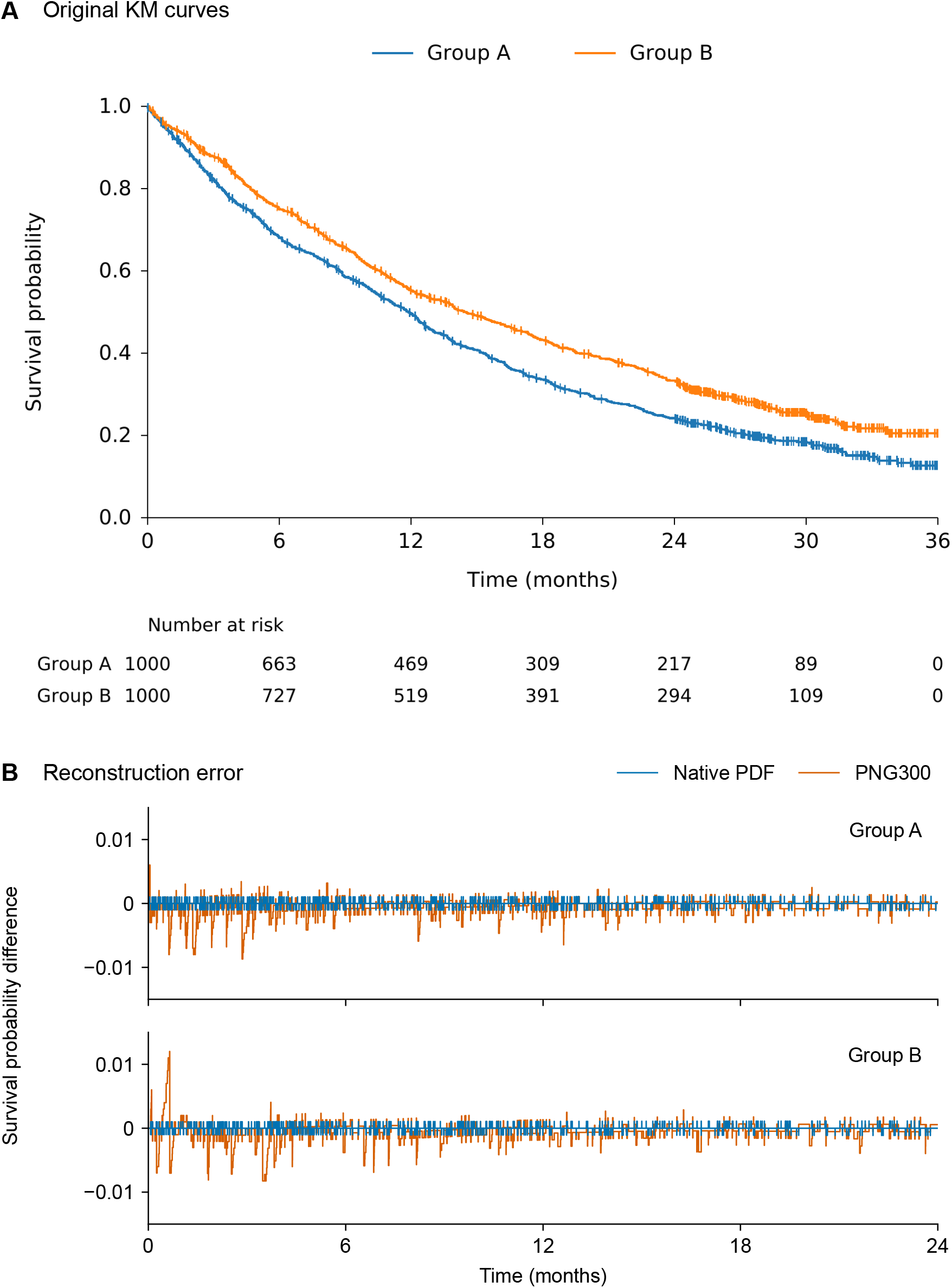
Example KM curves and reconstruction errors. (A) Original KM curves for dataset S3 D03 (1,000 participants per group). (B) Differences between reconstructed and reference survival probabilities over 24 months, by group and input format (native PDF or PNG300).

**Table 2.** Reconstruction accuracy for native PDFs and PNG300 images across 64 paired datasets. Values are mean absolute errors. Differences are PDF minus PNG300 with 95% stratified paired-bootstrap confidence intervals; negative values favor PDF. TAAE and RMST differences were evaluated over 24 months, while Cox log-HRs used the full follow-up.

| Outcome (unit) | Native PDF | PNG300 | PDF minus PNG300<br>mean difference [95% CI] |
|---|---|---|---|
| TAAE (pp) | 2.91 × 10<sup>−5</sup> | 0.1047 | −0.1047 [−0.1194, −0.0909] |
| Absolute error in the RMST difference (months) | 1.46 × 10<sup>−6</sup> | 0.00844 | −0.0084 [−0.0112, −0.0061] |
| Absolute Cox log-HR error | 4.51 × 10<sup>−6</sup> | 0.00267 | −0.0027 [−0.0036, −0.0019] |

Native PDFs also produced smaller errors in treatment-effect estimates than PNG300 images. Across the 64 datasets, mean absolute errors with PNG300 were 0.00844 months for the between-group 24-month RMST difference and 0.00267 for the Cox log-hazard ratio. The corresponding mean PDF-minus-PNG300 differences were −0.0084 months (95% CI,−0.0112 to −0.0061) and −0.0027 (95% CI, −0.0036 to −0.0019), respectively (Table 2).

Across 24 repeated-run comparisons per format, the mean time-averaged difference between first and repeated curves was 2.05 × 10^−5^ pp for native PDF and 0.127 pp for PNG300. These describe agreement between runs, rather than accuracy against reference records (S2 Table in S1 Text). In exploratory uncertainty analyses, the mean absolute relative error in the Cox standard error was 0.14% for PNG300 and negligible for native PDF (S10 Table in S1 Text).

### 3.2 Effects of figure characteristics

Removing censoring marks affected KM curve accuracy and agreement in event counts differently. For native PDFs, TAAE barely changed (difference, −5.9 × 10^−6^; 95% CI, −1.7 × 10^−5^ to 6.7 × 10^−6^), but reconstructed event totals matched the original records in both groups in only 28 of 32 datasets without marks, compared with all 32 when marks were present. Cox log-HR errors also increased for native PDFs (S1 Text). For PNG300 images, removing marks increased mean TAAE by 0.0227 (95% CI, 0.0046 to 0.0410; Fig 4).

**Fig 4.**
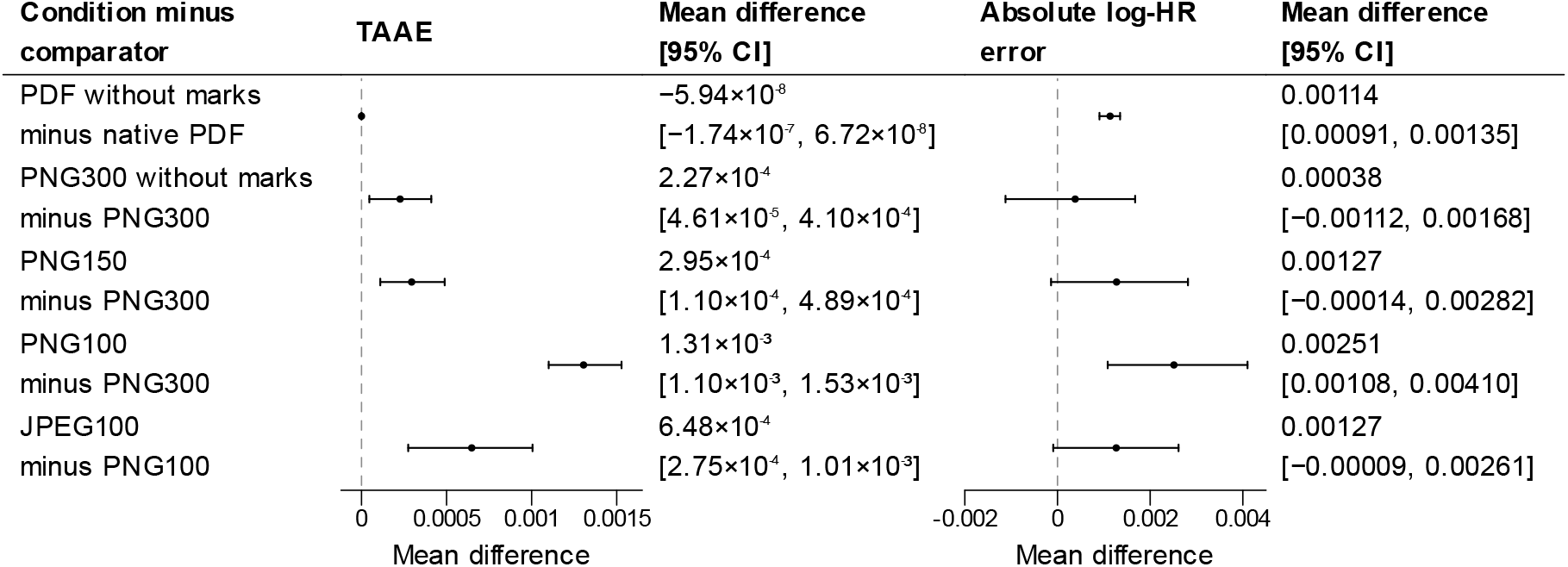
Effects of censoring marks, resolution, and compression in the 32-dataset exploratory subset. Points and intervals show mean paired differences and 95% scenario-stratified bootstrap confidence intervals. TAAE differences are shown on the survival-probability scale. Positive differences indicate larger errors for the modified input. Baseline outputs were reused from the primary experiment.

The post hoc analysis showed that small KM curve errors did not guarantee accurate event counts, even when both were evaluated after month 24. For native PDFs without censoring marks, event counts were correct through month 24 in all 32 datasets, but four datasets had one extra event after month 24. All four errors occurred within the later follow-up period evaluated, yet mean curve errors remained very small with and without marks (1.41 × 10^−5^ and 1.12 × 10^−5^ pp, respectively; S12 Table in S1 Text).

Lower image resolution and JPEG compression both increased KM reconstruction error. Compared with PNG300, reducing resolution to 150 dpi increased mean TAAE by 0.0295 (95% CI, 0.0110 to 0.0489), while reducing it to 100 dpi increased TAAE by 0.1305 (95% CI, 0.1100 to 0.1528). Converting PNG100 to quality-75 JPEG further increased mean TAAE by 0.0648 (95% CI, 0.0275 to 0.1005; Fig 4).

Saving a PNG image as a PDF did not restore the high reconstruction accuracy of native vector PDFs. Across the same 32 datasets, mean TAAE was 0.104 pp for image-only PDFs, compared with 3.40 × 10^−5^ for native vector PDFs and 0.0891 for PNG300 images. The mean paired difference between image-only PDFs and PNG300 was 0.0154 (95% CI, -0.0017 to 0.0321; S1 Text; S2 Figure). These findings support the interpretation that readable vector information contributes to the reconstruction advantage of native PDFs, beyond the PDF file format itself.

Native PDFs remained more accurate than PNG300 images when the KM figures were redrawn using survminer. Across all 16 datasets, native PDFs had lower TAAE, with mean errors of 0.000926 pp compared with 0.141 pp for PNG300. These findings suggest that the PDF advantage was not limited to the original plotting style, although the plotting style and execution period changed together (S1 Text; S5 Table; S1 Figure).

### 3.3 Exploratory comparison with an automated digitization–Guyot workflow

The LLM workflow produced smaller mean curve and treatment-effect errors than the evaluated automated digitization–Guyot workflow on the same PNG300 figures. Mean TAAE was 0.1047 pp for the LLM versus 0.1537 pp for the digitization–Guyot workflow (mean difference, 0.0490; 95% CI, 0.0342 to 0.0643). Mean absolute errors in the RMST difference were 0.0084 versus 0.0145 months, and those in the Cox log-HR were 0.0027 versus 0.0073, respectively (Table 3). These differences apply to the two implemented workflows, including their different use of censoring marks and opportunities for revision.

**Table 3.**
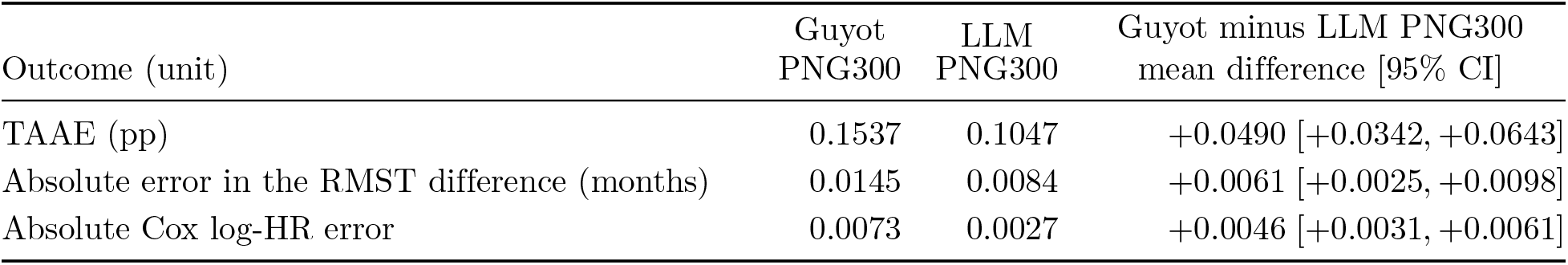
Exploratory comparison of the LLM and automated Guyot reconstruction using the same 64 PNG300 figures. Values are mean absolute errors relative to the original observed records. Differences are Guyot minus LLM PNG300; positive values favor the LLM. Approximate 95% confidence intervals (CIs) were obtained from 10,000 paired bootstrap resamples, sampling 16 datasets with replacement within each of four simulation scenarios. TAAE is time-averaged absolute KM error over 24 months, averaged across both arms. RMST differences (group B minus group A) were assessed at 24 months, while Cox log-hazard ratios used the full observed follow-up. pp, percentage points.

Although the LLM had lower average KM curve error, Guyot produced smaller errors at the time points where reconstructed curves differed most from the reference. Across the 64 datasets, mean maximum errors were 1.61 versus 1.90 pp in group A and 1.36 versus 1.75 pp in group B (Guyot versus LLM). These are averages of dataset-level maxima. This measure is sensitive to small horizontal shifts in KM steps, including numerical timing differences, so it should be interpreted alongside integrated errors (S1 Text).

In a post hoc comparison within the Guyot workflow, replacing digitized PNG300 coordinates with exact KM coordinates reduced mean curve error but increased mean errors in the RMST difference and Cox log-HR (S8 Table in S1 Text). Dataset-level diagnostics identified reduced cancellation between arm-specific RMST errors and changes in later event and censoring allocations (S1 Text).

## 4 Discussion

Preserving vector information improved KM reconstruction in this paired simulation study using a single tool-enabled language model. Native PDFs yielded lower curve errors than matched PNG300 inputs in every completed pair. However, the average PNG errors in RMST contrasts and Cox log-hazard ratios were small. The study establishes a difference in numerical fidelity within the evaluated workflow, without establishing clinically consequential changes in treatment comparisons or meta-analytic conclusions.

The saved programs provide a plausible explanation for the format difference: native-PDF reconstructions extracted stored curve paths, whereas PNG reconstructions estimated coordinates from pixels. An unchanged PNG embedded in a PDF did not recover the accuracy of the vector source. Earlier work demonstrated direct coordinate extraction from vector scientific figures, and RESOLVE-IPD applies related ideas to survival reconstruction [17, 30]. Our paired experiment quantifies the combined effect of information preservation and the agent’s response to that information. It does not establish that an LLM is necessary for accurate vector reconstruction. Nor should its performance be assumed to apply to a chat interface that cannot inspect vector objects or execute code.

Our findings complement the clinical validation by Everest et al., who found that RMST reconstruction accuracy was generally not associated with figure type across different published trials [18]. Their comparison and ours differ in design and outcome, and the small RMST discrepancies observed here do not establish a substantive contradiction. Assessing practical importance requires attention to the intended estimand, its uncertainty, and the distribution of errors rather than the frequency with which one format has the smaller error.

Accurate KM curves do not necessarily imply accurate recovery of individual event records. In the post hoc analysis, all four native-PDF event-count discrepancies occurred after month 24, outside the primary curve-error evaluation window. However, KM curve errors remained very small when evaluated over the same supported later follow-up, indicating that the discrepancy could not be explained by different evaluation windows alone. These findings support checking event totals and numbers at risk alongside KM curve agreement when validating reconstructed records.

The conventional comparison also requires qualification. The Guyot analyses showed that closer KM curve agreement did not always lead to more accurate treatment estimates. Although exact coordinates reduced curve error, mean RMST-contrast and Cox log-HR errors increased. Dataset-level diagnostics suggested that these differences reflected reduced cancellation between arm-specific RMST errors and changes in late event and censoring allocations. For some arms, coordinate substitution also changed the final risk-table interval constraints (S8 Table in S1 Text). These results describe how the Guyot workflow responds to different inputs rather than separate contributions of digitization and reconstruction error. The LLM–Guyot comparison likewise reflects differences between the two implemented workflows, including their use of censoring marks and opportunities for revision. Future comparisons should use validated implementations and explicitly matched information where the aim is to isolate algorithmic performance.

For practice, researchers should preserve original vector figures when available and verify that their extraction workflow can use the encoded information. Saving a screenshot as a PDF does not restore vector coordinates. Journals and repositories can support reuse by retaining vector graphics and reporting risk counts and censoring information [31,32]. This recommendation concerns preserved content, rather than a preference for a filename extension. Reconstructed outputs still require checks appropriate to the planned survival analysis.

The study has several limitations. First, the simulations used exponential or piecewise-exponential event times, one baseline event rate, and one dropout rate. Crossing hazards broadened the design, but other patterns, including Weibull hazards, were not evaluated. Complete risk tables, clean two-arm figures, and limited random censoring before month 24 also provided favorable conditions. Published figures may contain incomplete risk information, overlapping elements, and different censoring patterns [33–35]. The number of plotted groups and the complexity of figure legends and labels may also affect reconstruction accuracy. The primary evaluation omitted later follow-up, where administrative censoring accumulated. Second, only one model, reasoning setting, and tool environment were evaluated. Replacement runs and unenforced revision limits mean that the results describe the implemented workflow, rather than first-attempt performance or performance under a uniform revision budget. Third, 16 datasets per scenario and limited repeat runs provide incomplete characterization of scenario-specific and model-run variability. Later exploratory experiments reused earlier baseline outputs, so execution period may also contribute to those contrasts.

Further work should prioritize published-figure validation, comparable reconstruction information and resource limits, and repeated runs under documented model versions. Where original clinical records are unavailable, agreement with reported summaries can assess consistency but cannot establish participant-level truth. Studies with original records can assess downstream estimands directly. More varied simulations would help identify when reconstruction errors become consequential, rather than merely demonstrating smaller errors in additional favorable examples.

## 5 Conclusion

Native vector figures supported more accurate reconstruction than matched images in the evaluated language-model workflow, while average differences in treatment estimates remained small. Preserving figure information and checking reconstructed outcomes are both relevant to reliable survival evidence synthesis. The results require further validation under more varied figure conditions and consistently enforced reconstruction procedures.

## Supporting information

**S1 Text. Supplementary methods and results.** Supplementary methods, protocol deviations, sensitivity analyses, and exploratory comparisons.

## Declarations

### Author contributions

Shiyin Li: Conceptualization, Data curation, Formal analysis, Investigation, Methodology, Project administration, Software, Validation, Visualization, Writing – original draft, Writing – review & editing. Xing Xing: Writing – review & editing. Bulun Te: Writing – review & editing. Han Han: Writing – review & editing. Haoyu Ji: Writing – review & editing. Yulin Yu: Writing – review & editing. Lianne Siegel: Writing – review & editing. Lifeng Lin: Conceptualization, Funding acquisition, Supervision, Writing – review & editing.

### Ethics

This study used exclusively simulated data and did not involve human participants. Therefore, ethics approval and informed consent were not required.

### AI assistance

The authors used OpenAI Codex to assist with analysis code development and language editing to improve manuscript clarity and readability. All AI-assisted outputs were reviewed and verified by the authors, who take full responsibility for the final content. The language model evaluated for KM reconstruction is described separately in Methods.

## Data Availability

All data used in this study were simulated. The project repository is https://github.com/FionaShiyinLi/km-curve-reconstruction-study. The reproducibility materials comprise reference records, input figures, reconstructed records, dataset-level results, reconstruction instructions, random seeds, and generation, reconstruction, and analysis code.

## References

1. Ohmann C, Moher D, Siebert M, Motschall E, Naudet F. Status, use and impact of sharing individual participant data from clinical trials: a scoping review. BMJ Open. 2021;11(8):e049228. doi:10.1136/bmjopen-2021-049228.

2. Zhao Y, Sun H, Ding Y, Xu Y. KM-GPT: an automated pipeline for reconstructing individual patient data from Kaplan–Meier plots; 2025. Preprint. arXiv:2509.18141, version 1. Available from: https://arxiv.org/abs/2509.18141v1. doi:10.48550/arXiv.2509.18141.

3. Jiang J, Gao C, Rasmussen E, Xie SZ, Sun J. KMGen: a skill-based approach for synthetic individual patient data generation. In: Proceedings of the 11th Machine Learning for Healthcare Conference. vol. 340 of Proceedings of Machine Learning Research. PMLR; 2026. p. 755–814. Available from: https://proceedings.mlr.press/v340/jiang26a.html.

4. Kaplan EL, Meier P. Nonparametric estimation from incomplete observations. J Am Stat Assoc. 1958;53(282):457–81. doi:10.1080/01621459.1958.10501452.

5. Royle KL, Meads D, Visser-Rogers JK, White IR, Cairns DA. How is overall survival assessed in randomised clinical trials in cancer and are subsequent treatment lines considered? A systematic review. Trials. 2023;24:708. doi:10.1186/s13063-023-07730-1.

6. Tierney JF, Stewart LA, Ghersi D, Burdett S, Sydes MR. Practical methods for incorporating summary time-to-event data into meta-analysis. Trials. 2007;8:16. doi:10.1186/1745-6215-8-16.

7. Guyot P, Ades AE, Ouwens MJNM, Welton NJ. Enhanced secondary analysis of survival data: reconstructing the data from published Kaplan–Meier survival curves. BMC Med Res Methodol. 2012;12:9. doi:10.1186/1471-2288-12-9.

8. Wei Y, Royston P. Reconstructing time-to-event data from published Kaplan–Meier curves. Stata J. 2017;17(4):786–802. doi:10.1177/1536867X1801700402.

9. Rogula B, Lozano-Ortega G, Johnston KM. A method for reconstructing individual patient data from Kaplan–Meier survival curves that incorporate marked censoring times. MDM Policy Pract. 2022;7(1):23814683221077643. doi:10.1177/23814683221077643.

10. Titman AC. Using quadratic programming to reconstruct data from published survival and competing risks analyses. Stat Med. 2026;45(6–7):e70474. doi:10.1002/sim.70474.

11. Zhang JZ, Rios JD, Pechlivanoglou T, Yang A, Zhang Q, Deris D, et al. SurvdigitizeR: an algorithm for automated survival curve digitization. BMC Med Res Methodol. 2024;24:147. doi:10.1186/s12874-024-02273-8.

12. Yao S, Zhao J, Yu D, Du N, Shafran I, Narasimhan K, et al. ReAct: Synergizing reasoning and acting in language models. In: International Conference on Learning Representations; 2023. Available from: https://arxiv.org/abs/2210.03629.

13. Surís D, Menon S, Vondrick C. ViperGPT: Visual inference via Python execution for reasoning. In: Proceedings of the IEEE/CVF International Conference on Computer Vision; 2023. p. 11888–98. Available from: https://openaccess.thecvf.com/content/ICCV2023/html/Suris_ViperGPT_Visual_Inference_via_Python_Execution_for_Reasoning_ICCV_2023_paper.html.

14. Adobe Systems Incorporated. PDF Reference: Adobe Portable Document Format, Version 1.7; 2006. Sections 2.1.2–2.1.4 and 6.5; accessed 25 September 2026. Available from: https://opensource.adobe.com/dc-acrobat-sdk-docs/pdfstandards/pdfreference1.7old.pdf.

15. Praczyk PA, Nogueras-Iso J. Automatic extraction of figures from scientific publications in high-energy physics. Inf Technol Libr. 2013;32(4):25–52. doi:10.6017/ital.v32i4.3670.

16. Kim J, Chalise P, He J. Reconstructing patient level survival data from published Kaplan–Meier curves. Contemp Clin Trials Commun. 2025;47:101542. doi:10.1016/j.conctc.2025.101542.

17. Lang L, Zhao Y, Gao Q, Xu Y. RESOLVE-IPD: high-fidelity individual patient data reconstruction and uncertainty-aware subgroup meta-analysis; 2026. Preprint; submitted 3 November 2025, revised 21 January 2026. arXiv:2511.01785, version 2. Available from: https://arxiv.org/abs/2511.01785v2. doi:10.48550/arXiv.2511.01785.

18. Everest L, Blommaert S, Tu D, Pater JL, Hay A, Cheung MC, et al. Validating Restricted Mean Survival Time Estimates From Reconstructed Kaplan–Meier Data Against Original Trial Individual Patient Data From Trials Conducted by the Canadian Cancer Trials Group. Value Health. 2022;25(7):1157–64. doi:10.1016/j.jval.2021.12.004.

19. Morris TP, White IR, Crowther MJ. Using simulation studies to evaluate statistical methods. Stat Med. 2019;38(11):2074–102. doi:10.1002/sim.8086.

20. Wan X, Peng L, Li Y. A Review and Comparison of Methods for Recreating Individual Patient Data from Published Kaplan-Meier Survival Curves for Economic Evaluations: A Simulation Study. PLoS One. 2015;10(3):e0121353. doi:10.1371/journal.pone.0121353.

21. Rahman R, Fell G, Ventz S, Arfé A, Vanderbeek AM, Trippa L, et al. Deviation from the Proportional Hazards Assumption in Randomized Phase 3 Clinical Trials in Oncology: Prevalence, Associated Factors, and Implications. Clin Cancer Res. 2019;25(21):6339–45. doi:10.1158/1078-0432.CCR-18-3999.

22. L’Ecuyer P, Simard R, Chen EJ, Kelton WD. An object-oriented random-number package with many long streams and substreams. Oper Res. 2002;50(6):1073–5. doi:10.1287/opre.50.6.1073.358.

23. Royston P, Parmar MKB. Restricted mean survival time: an alternative to the hazard ratio for the design and analysis of randomized trials with a time-to-event outcome. BMC Med Res Methodol. 2013;13:152. doi:10.1186/1471-2288-13-152.

24. Uno H, Claggett B, Tian L, Inoue E, Gallo P, Miyata T, et al. Moving Beyond the Hazard Ratio in Quantifying the Between-Group Difference in Survival Analysis. J Clin Oncol. 2014;32(22):2380–5. doi:10.1200/JCO.2014.55.2208.

25. Cox DR. Regression models and life-tables. J R Stat Soc Series B Stat Methodol. 1972;34(2):187–202. doi:10.1111/j.2517-6161.1972.tb00899.x.

26. Efron B. The Efficiency of Cox’s Likelihood Function for Censored Data. J Am Stat Assoc. 1977;72(359):557–65. doi:10.1080/01621459.1977.10480613.

27. Hertz-Picciotto I, Rockhill B. Validity and efficiency of approximation methods for tied survival times in Cox regression. Biometrics. 1997;53(3):1151–6. doi:10.2307/2533573.

28. DiCiccio TJ, Efron B. Bootstrap confidence intervals. Stat Sci. 1996;11(3):189–228. doi:10.1214/ss/1032280214.

29. Kassambara A, Kosinski M, Biecek P. survminer: Drawing Survival Curves using ggplot2; 2026. R package version 0.5.2. doi:10.32614/CRAN.package.survminer.

30. Hartgerink C, Murray-Rust P. Extracting data from vector figures in scholarly articles; 2018. Originally posted 2017; revised 12 March 2018; cited 19 October 2026. arXiv:1709.02261v3 [Preprint]. Available from: https://arxiv.org/abs/1709.02261v3.

31. Pocock SJ, Clayton TC, Altman DG. Survival plots of time-to-event outcomes in clinical trials: good practice and pitfalls. Lancet. 2002;359(9318):1686–9. doi:10.1016/S0140-6736(02)08594-X.

32. Morris TP, Jarvis CI, Cragg W, Phillips PPJ, Choodari-Oskooei B, Sydes MR. Proposals on Kaplan–Meier plots in medical research and a survey of stakeholder views: KMunicate. BMJ Open. 2019;9(9):e030215. doi:10.1136/bmjopen-2019-030215.

33. Sherry AD, Msaouel P, Kouzy R, Abi Jaoude J, Lin TA, Taniguchi CM, et al. Lost in the plot: missing visual elements in Kaplan–Meier plots of phase III oncology trials. Oncologist. 2024;29(7):547–50. doi:10.1093/oncolo/oyae067.

34. Vervölgyi E, Kromp M, Skipka G, Bender R, Kaiser T. Reporting of loss to follow-up information in randomised controlled trials with time-to-event outcomes: a literature survey. BMC Med Res Methodol. 2011;11:130. doi:10.1186/1471-2288-11-130.

35. Rosen K, Prasad V, Chen EY. Censored patients in Kaplan–Meier plots of cancer drugs: An empirical analysis of data sharing. Eur J Cancer. 2020;141:152–61. doi:10.1016/j.ejca.2020.09.031.

